# SARS-CoV-2 Omicron sublineage BA.2 replaces BA.1.1: genomic surveillance in Japan from September 2021 to March 2022

**DOI:** 10.1101/2022.04.05.22273483

**Authors:** Yosuke Hirotsu, Makoto Maejima, Masahiro Shibusawa, Yume Natori, Yuki Nagakubo, Kazuhiro Hosaka, Hitomi Sueki, Hitoshi Mochizuki, Toshiharu Tsutsui, Yumiko Kakizaki, Yoshihiro Miyashita, Masao Omata

## Abstract

**Objective:** The new emerging Omicron strain of severe acute respiratory syndrome coronavirus 2 (SARS-CoV-2) is currently spreading worldwide. We aimed to analyze the genomic evolution of the shifting Omicron virus subtypes.

**Methods:** The study included 1,297 individuals diagnosed as SARS-CoV-2 positive by PCR test or antigen quantification test from September 2021 to March 2022. Samples were analyzed by whole genome sequencing analysis (n=489) or TaqMan assay (n=808).

**Results:** After the outbreak of the SARS-CoV-2 Delta strain, the Omicron strain spread rapidly in Yamanashi, Japan. BA.1.1 was the predominant sublineage of the Omicron strain from January to mid-February 2022, but the number of cases of sublineage BA.2 began to increase after mid-February, and this sublineage was shown to have replaced BA.1.1 by the end of March 2022. We observed higher viral and antigen levels of sublineage BA.2 than of sublineage BA.1.1 in nasopharyngeal swab samples. However, no difference in viral load by patient age was apparent between sublineages BA.1.1 and BA.2.

**Conclusions:** A transition from sublineage BA.1.1 to sublineage BA.2 was clearly observed over approximately one month. Omicron sublineage BA.2 was found to be more transmissible owing to its higher viral load regardless of patient age.

## Introduction

Since the discovery of severe acute respiratory syndrome coronavirus 2 (SARS-CoV-2) at the end of 2019, large numbers of infections and deaths have been reported. The Omicron (B.1.1.529) strain of SARS-CoV-2 was first identified in South Africa, and infection with Omicron has been confirmed in 169 countries to date [1, 2]. World Health Organization designated the Omicron strain as a variant of concern at the end of November 2021 [1]. Now several Omicron strain sublineages, such as BA.1, BA.1.1, BA.2, and BA.3, have been described.

The Omicron strain has multiple spike protein mutations compared with other variants of concern, such as the Alpha and Delta strains [2]. Consequently, there is concern that serum antibody activity against the Omicron strain in vaccinated or convalescent persons will be weaker than that against previous SARS-CoV-2 strains [3, 4]. In addition, for some antibody therapies, the level of neutralizing activity was shown to differ between Omicron sublineages BA.1 and BA.2 [5, 6]. Omicron strains are considered to be highly transmissible but have a relatively lower critical illness risk [7-10]. In many countries, Omicron strains are rapidly increasing in prevalence and affecting medical and social activities. Because the characteristics of infectivity and treatment response differ among Omicron sublineages, it is important to understand the evolutionary process in real time.

In this study, we conducted whole genome sequencing analyses and TaqMan assays of SARS-CoV-2 on nasopharyngeal swab samples collected from 1,297 patients in Japan from September 2021 to March 2022 to investigate the evolution of SARS-CoV-2 strains.

## Methods

### SARS-CoV-2 diagnostic testing

Multiple molecular diagnostic testing platforms, including COVID-19 reverse transcription-PCR performed in accordance with the protocol developed by the National Institute of Infectious Diseases in Japan [11], the FilmArray Respiratory Panel 2.1 test performed with the FilmArray Torch system (bioMérieux, Marcy-l’Etoile, France) [12], the Xpert Xpress SARS-CoV-2 test performed with Cepheid GeneXpert (Cepheid, Sunnyvale, CA, USA) [13], and the Lumipulse antigen test performed with the LUMIPULSE G600II system (Fujirebio, Inc., Tokyo, Japan) were used for this study [14, 15]. All tests were conducted on material obtained from nasopharyngeal swabs immersed in viral transport media (Copan, Murrieta, CA, USA).

### Quantitative reverse transcription-PCR (RT-qPCR)

To detect SARS-CoV-2, we performed one-step RT-qPCR amplifying the nucleocapsid (N) gene of SARS-CoV-2, as we described previously [16]. The human ribonuclease P protein subunit p30 (*RPP30*) gene was used as the internal positive control (Integrated DNA Technologies, Coralville, IA, USA) [16].

The RT-qPCR assays were performed on a StepOnePlus Real-Time PCR System (Thermo Fisher Scientific) with the following cycling conditions: reverse transcription at 50 °C for 5 min, inactivation of reverse transcription at 95 °C for 20 s, and denature, annealing and extension at 45 cycles of 95 °C for 3 s, 60 °C for 30 s. The threshold was set at 0.2. In accordance with the national protocol (version 2.9.1) [11], samples were assessed as positive if a visible amplification plot was observed and as negative if no amplification was observed.

### SARS-CoV-2 genome analysis

Whole genome sequencing analysis was conducted in accordance with a previously described method on 489 nasopharyngeal swabs collected from patients with coronavirus disease 2019 (COVID-19) from September 2021 to March 2022. In brief, SARS-CoV-2 genomic RNA was reverse-transcribed into cDNA and amplified using the Ion AmpliSeq SARS-CoV-2 Research Panel or Ion AmpliSeq SARS-CoV-2 Insight Research Assay (Thermo Fisher Scientific, Waltham, MA, USA) on the Ion Torrent Genexus System in accordance with the manufacturer’s instructions [17-19]. Sequencing reads were processed, and their quality was assessed using Genexus Software with SARS-CoV-2 plugins. The sequencing reads were then mapped and aligned using the torrent mapping alignment program. After initial mapping, a variant call was performed using the Torrent Variant Caller. The COVID19AnnotateSnpEff plugin was used to annotate the variants. Assembly was performed using the Iterative Refinement Meta-Assembler [20].

The viral clade and lineage classifications were conducted using Nextstrain [21] and Phylogenetic Assignment of Named Global Outbreak Lineages (PANGOLIN) [22]. Sequence data were deposited in the Global Initiative on Sharing Avian Influenza Data (GISAID) EpiCoV database [23].

### TaqMan assay

We used the pre-designed TaqMan SARS-CoV-2 Mutation Panel for detecting SARS-CoV-2 spike Δ69–70, G339D, L452R, and/or Q493R (Thermo Fisher Scientific) using 808 SARS-CoV-2-positive samples (in submission). The TaqMan MGB probe for the wild-type allele was labelled with VIC dye, and the probe for the variant allele was labelled with FAM dye. This TaqMan probe system detected both wild-type and variant sequences of SARS-CoV-2. TaqPath 1-Step RT-qPCR Master Mix CG was used as master mix. Real-time PCR was conducted on a Step-One Plus Real Time PCR System (Thermo Fisher Scientific).

## Results

### Transition of SARS-CoV-2 strain prevalence

To determine the viral lineage of SARS-CoV-2, we performed whole genome sequencing analyses or TaqMan assays using SARS-CoV-2-positive samples (n = 1,297) collected consecutively in Yamanashi, Japan from September 2021 to March 2022 (Figure 1A). During this period, we identified Delta strain (n = 159) and Omicron strain (n = 1,139). After the first case of Omicron was identified in January 2022, Omicron rapidly replaced Delta as the prevalent strain of SARS-CoV-2 (Figure 1A).

**Figure 1.**
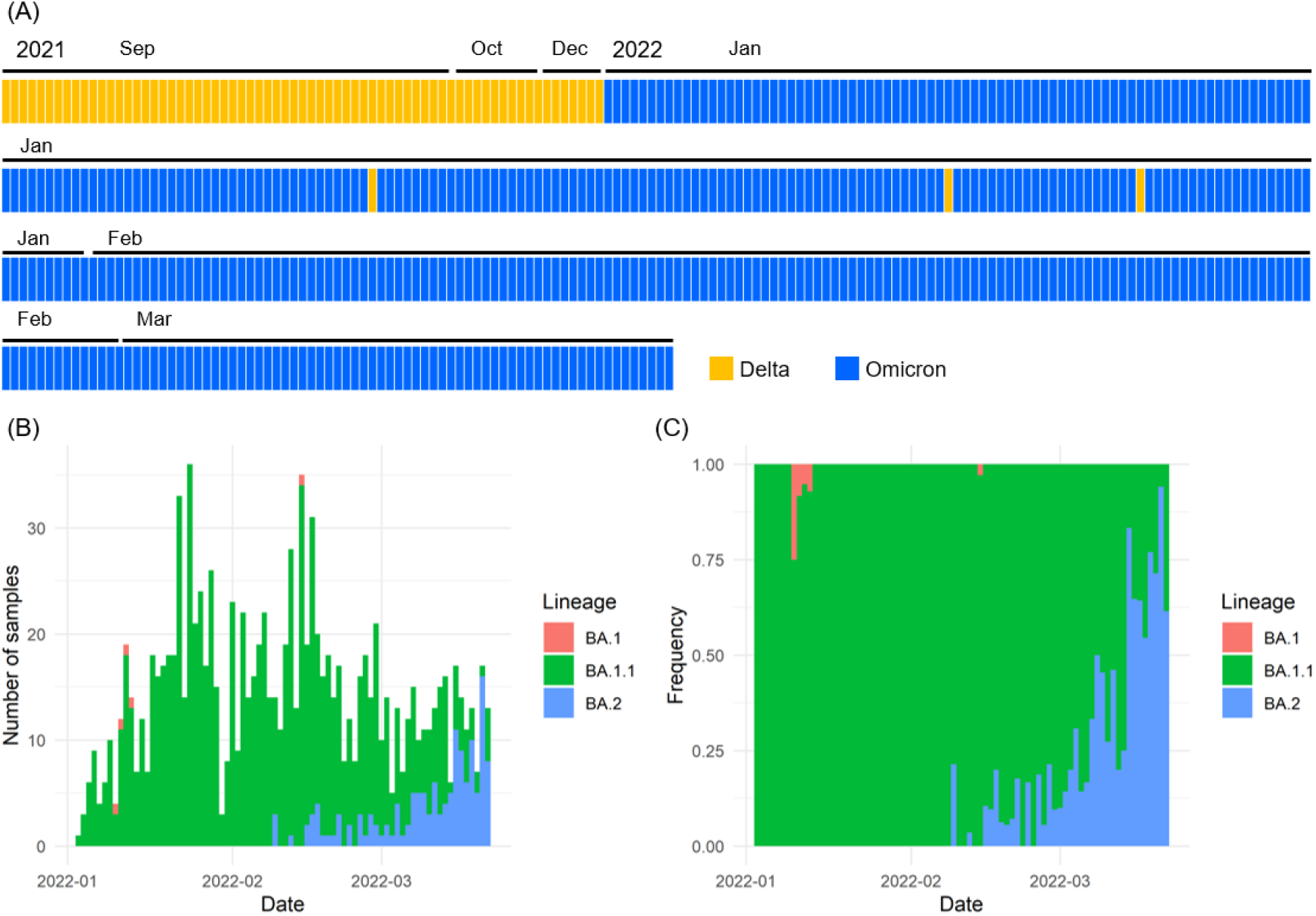
Changes in Omicron strain prevalence. **(A)** SARS-CoV-2 strains identified from September 2021 to March 2022. Orange boxes indicate Delta strains, and blue boxes indicate Omicron strains. **(B, C)** Sublineage of Omicron strains detected from January 2022 to March 2022, indicated by BA.1 (pink), BA.1.1 (green), and BA.2 (blue). The number of samples detected per day (B) and the frequency of detection (C) are shown.

### Changes in Omicron sublineages

The whole genome sequencing data were analyzed using PANGOLIN (version 3.1.20), and BA.1 (n = 5), BA.1.1 (n = 992), and BA.2 (n = 142) were identified as sublineages of Omicron (Figure 1B). Sublineage BA.1.1 was the dominant sublineage of Omicron from January to mid-February 2022; however, the incidence of sublineage BA.2 increased from mid-February 2022 onward, with this sublineage becoming dominant by the end of March (Figure 1B and 1C). The average frequency for the seven-day period from March 8 to March 14 was 62.2% (51/82) for sublineage BA.1.1 and 37.8% (31/82) for sublineage BA.2, whereas from March 15 to March 21 it was 29.3% (27/92) for sublineage BA.1.1 and 70.7% (65/92) for sublineage BA.2. These results indicate an extremely rapid replacement of sublineage BA.1.1 by sublineage BA.2 and a higher transmissibility of sublineage BA.2 compared with sublineage BA.1.1.

### Viral load of Omicron sublineages

To investigate the underlying factors for the high transmissibility of Omicron sublineage BA.2, we performed an RT-qPCR analysis of the viral load in the nasopharyngeal swabs collected from patients infected with sublineage BA.1.1 (n = 748) or sublineage BA.2 (n = 118). The median viral load (log_10_ copies/mL) was 5.7 (range: 0.2–7.9) for sublineage BA.1.1 versus 6.4 (range: 0.3–8.2) for sublineage BA.2 (Figure 2A). The median Ct value for sublineage BA.1.1 was 19 (range: 11–38) versus 17 (range: 10–38) for sublineage BA.2 (Figure 2B). There are significant differences in the viral load between cases of sublineage BA.1.1 and sublineage BA.2 (Figure 2A, *p* = 4.8×10^−4^, Student’s *t*-test) and Ct value (Figure 2B, *p* = 1.6×10^−3^, Student’s *t*-test).

**Figure 2.**
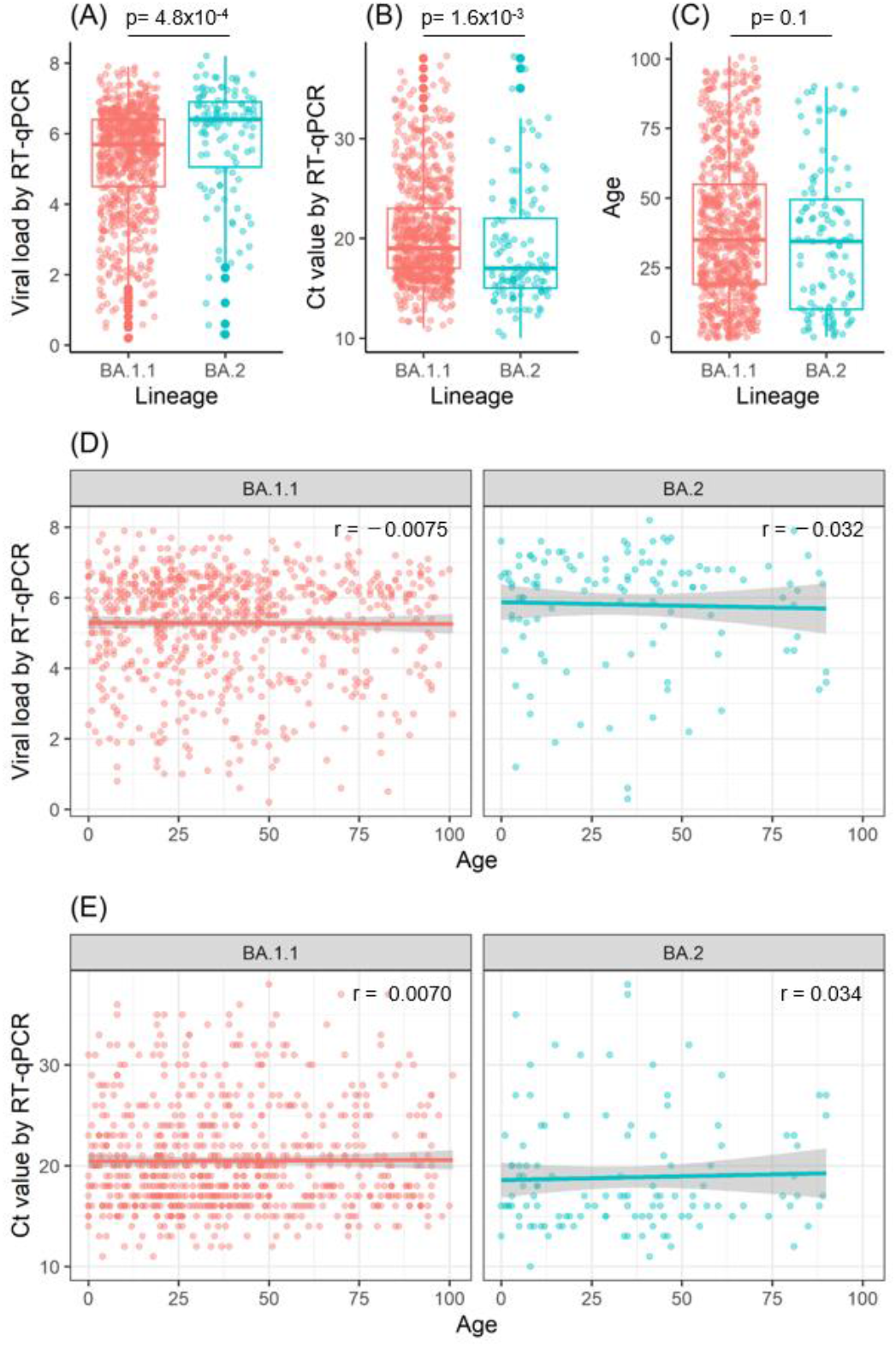
Viral load and age of infected patients for sublineages BA.1. and BA.2. **(A, B)** The viral load and Ct values in Omicron sublineages BA.1.1 (n = 748) and BA.2 (n = 118) were analyzed by RT-qPCR. Box plots show the viral load (A) and Ct values (B) in BA.1.1 and BA.2. **(C)** Box plot shows the age of patients infected with sublineage BA.1.1 or BA.2. **(D, E)** Relationship between patient age and viral load (D) or Ct value (E). Pearson’s correlation coefficient (r) is noted in the figures. The gray background of the regression line indicates the 95% confidence interval.

Of the 866 samples, 827 were tested for SARS-CoV-2 antigen, and the percentage of samples with high antigen levels was examined. The percentage of specimens with antigen levels of >5000 pg/mL was 57.3% (386/711) for sublineage BA.1.1 and 69% (80/116) for sublineage BA.2, indicating that sublineage BA.2 had higher antigen levels (*p* = 0.004, chi-squared test). However, the median age of infected patients was not significantly different between these sublineages (35 years [range: 0–101 years] for BA.1.1 vs. 34.5 years [range: 0–90 years] for BA.2; *p* = 0.1, Student’s *t*-test) (Figure 2C). These results indicate that the viral load in nasopharyngeal swabs is higher for sublineage BA.2 than for sublineage BA.1.1 and that sublineage BA.2 is more contagious.

We next examined whether the viral load varied with patient age. There was no apparent correlation between patient age and viral load or Ct value for either sublineage BA.1.1 or BA.2 (Figure 3D and 3E). The Pearson’s correlation coefficients for sublineage BA.1.1 were r = −0.0075 (*p* = 0.84) for patient age and viral load and r = 0.0070 (*p* = 0.85) for patient age and Ct value, and those for sublineage BA.2 were r = −0.032 (*p* = 0.73) for patient age and viral load and r = 0.034 (*p* = 0.71) for patient age and Ct value (Figures 2D and 2E). These results indicate that the viral load remained fairly high in Omicron-infected patients regardless of their age.

## Discussion

This study indicates that after the expansion of the SARS-CoV-2 Delta strain, a rapid spread of the Omicron strain occurred. Sublineage BA.1 was very minor in Japan when Omicron was first discovered. First, sublineage BA.1.1 expanded dominantly and was then gradually replaced by sublineage BA.2. The results of the present study show that the amount of viral load in the nasopharyngeal swab was higher for sublineage BA.2 than for sublineage BA.1.1. These epidemiological and viral characteristic results indicate that Omicron sublineage BA.2 is more transmissible than sublineage BA.1.1.

Previous reports showed that sublineage BA.2 has a lower Ct value (i.e., higher viral load) compared with sublineages BA.1 and BA.1.1 [24-26]. The findings of the present study are consistent with those reports, and this difference could be one reason for the higher infectivity of sublineage BA.2. In Denmark, England, India, the Philippines, and South Africa, where Omicron strains were predominantly sublineage BA.1 in the early stages of the outbreak, sublineage BA.2 later became predominant [27]. Furthermore, reinfection with sublineage BA.2 after infection with sublineage BA.1 can occur, although it is rare [28]. Therefore, there is concern that the prevalence of the BA.2 sublineage may increase in the future.

The relationship between age and SARS-CoV-2 viral load of other strains was shown previously [29-33]; however, no data on Omicron sublineages were reported. Previous studies suggested that the SARS-CoV-2 viral load tends to be higher in young children than in adults, whereas other data suggest that the viral load does not vary by age group [29-33]. In this study, no obvious differences in viral load by age group were observed for either the Omicron BA.1.1 or BA.2 sublineages. In general, viral load peaks in the early phase of infection and then gradually declines; hence, the timing of sampling relative to the onset of symptoms is an important factor [34]. Because the time between onset and sampling was not taken into account in the present study, our data are limited by sampling bias. However, our data are derived from random sampling, therefore these results are expected to better reflect real-world conditions. Although a high incidence of household COVID-19 infections stemming from young children has been reported [35], our results indicate that the Omicron strain retains a fairly high viral load across age groups, which may contribute to the high infectivity of the Omicron strain and its accelerated spread. These data provide insights for determining appropriate COVID-19 prevention and control measures for homes, schools, workplaces, and facilities for the elderly during the spread of Omicron strain viruses.

Recombinant variants may emerge in communities where SARS-CoV-2 strains with different genomic architecture are co-circulating [36]. Recently, a new hybrid strain (AY.4/BA.1 recombinant, EPI_ISL_10819657), which has the properties of both the Delta and Omicron strains, was reported in France [23]. This hybrid virus has been detected in several countries, including Belgium, Germany, Denmark, and the Netherlands. There is also evidence for co-infection and recombination events between Delta and Omicron strains in the same patient [37]. Recombinant viruses BA.1 and BA.2 (named XE) were also reported from the United Kingdom [38]. It is not yet fully understood whether these hybrids are highly infectious, pathogenic, or resistant to antibodies or therapy. Therefore, it is necessary to survey the hybrid viruses to see if there is likely to be an explosion of infections. Genomic epidemiological analysis of SARS-CoV-2 strains and variants should be continued to monitor future virus trends.

## Data Availability

All data produced in the present work are contained in the manuscript.

## Acknowledgements

We thank all medical and ancillary hospital staff for their support. We thank Katie Oakley, PhD, from Edanz (https://jp.edanz.com/ac) for editing a draft of this manuscript.

## References

1. World Health Organization. Classification of Omicron (B.1.1.529): SARS-CoV-2 Variant of Concern. 2021.

2. Julia LM, Ginger T, Alaa AL, Manar A, Marco C, Emily H, et al. outbreak.info.

3. Iketani S, Liu L, Guo Y, Liu L, Chan JFW, Huang Y, et al. Antibody evasion properties of SARS-CoV-2 Omicron sublineages. Nature. 2022. doi: 10.1038/s41586-022-04594-4.

4. Yu J, Collier A-rY, Rowe M, Mardas F, Ventura JD, Wan H, et al. Neutralization of the SARS-CoV-2 Omicron BA.1 and BA.2 Variants. New England Journal of Medicine. 2022. doi: 10.1056/NEJMc2201849.

5. Bruel T, Hadjadj J, Maes P, Planas D, Seve A, Staropoli I, et al. Serum neutralization of SARS-CoV-2 Omicron sublineages BA.1 and BA.2 in patients receiving monoclonal antibodies. Nature Medicine. 2022. doi: 10.1038/s41591-022-01792-5.

6. Takashita E, Kinoshita N, Yamayoshi S, Sakai-Tagawa Y, Fujisaki S, Ito M, et al. Efficacy of Antiviral Agents against the SARS-CoV-2 Omicron Subvariant BA.2. New England Journal of Medicine. 2022. doi: 10.1056/NEJMc2201933.

7. Wolter N, Jassat W, Walaza S, Welch R, Moultrie H, Groome M, et al. Early assessment of the clinical severity of the SARS-CoV-2 omicron variant in South Africa: a data linkage study. The Lancet. 2022;399(10323):437–46. doi: 10.1016/S0140-6736(22)00017-4.

8. Christensen PA, Olsen RJ, Long SW, Snehal R, Davis JJ, Saavedra MO, et al. Signals of significantly increased vaccine breakthrough, decreased hospitalization rates, and less severe disease in patients with COVID-19 caused by the Omicron variant of SARS-CoV-2 in Houston, Texas. The American Journal of Pathology. 2022. doi: 10.1016/j.ajpath.2022.01.007.

9. Veneti L, Bøås H Bråthen Kristoffersen A, Stålcrantz J, Bragstad K, Hungnes O, et al. Reduced risk of hospitalisation among reported COVID-19 cases infected with the SARS-CoV-2 Omicron BA.1 variant compared with the Delta variant, Norway, December 2021 to January 2022. Eurosurveillance. 2022;27(4):2200077. doi: doi:https://doi.org/10.2807/1560-7917.ES.2022.27.4.2200077.

10. Espenhain L, Funk T, Overvad M, Edslev SM, Fonager J, Ingham AC, et al. Epidemiological characterisation of the first 785 SARS-CoV-2 Omicron variant cases in Denmark, December 2021. Eurosurveillance. 2021;26(50):2101146. doi: doi:https://doi.org/10.2807/1560-7917.ES.2021.26.50.2101146.

11. Shirato K, Nao N, Katano H, Takayama I, Saito S, Kato F, et al. Development of Genetic Diagnostic Methods for Novel Coronavirus 2019 (nCoV-2019) in Japan. Jpn J Infect Dis. 2020;73(4):304–7 doi: 10.7883/yoken.JJID.2020.061. PubMed PMID: 32074516.

12. Hirotsu Y, Maejima M, Shibusawa M, Amemiya K, Nagakubo Y, Hosaka K, et al. Analysis of Covid-19 and non-Covid-19 viruses, including influenza viruses, to determine the influence of intensive preventive measures in Japan. J Clin Virol. 2020;129:104543. doi: 10.1016/j.jcv.2020.104543

13. Hirotsu Y, Maejima M, Shibusawa M, Natori Y, Nagakubo Y, Hosaka K, et al. Direct comparison of Xpert Xpress, FilmArray Respiratory Panel, Lumipulse antigen test, and RT-qPCR in 165 nasopharyngeal swabs. BMC Infectious Diseases. 2022;22(1):221. doi: 10.1186/s12879-022-07185-w.

14. Hirotsu Y, Maejima M, Shibusawa M, Amemiya K, Nagakubo Y, Hosaka K, et al. Prospective Study of 1,308 Nasopharyngeal Swabs from 1,033 Patients using the LUMIPULSE SARS-CoV-2 Antigen Test: Comparison with RT-qPCR. International Journal of Infectious Diseases. 2021. doi: https://doi.org/10.1016/j.ijid.2021.02.005.

15. Hirotsu Y, Maejima M, Shibusawa M, Nagakubo Y, Hosaka K, Amemiya K, et al. Comparison of Automated SARS-CoV-2 Antigen Test for COVID-19 Infection with Quantitative RT-PCR using 313 Nasopharyngeal Swabs Including from 7 Serially Followed Patients. International Journal of Infectious Diseases. 2020. doi: https://doi.org/10.1016/j.ijid.2020.08.029.

16. Hirotsu Y, Mochizuki H, Omata M. Double-quencher probes improve detection sensitivity toward Severe Acute Respiratory Syndrome Coronavirus 2 (SARS-CoV-2) in a reverse-transcription polymerase chain reaction (RT-PCR) assay J Virol Methods. 2020;284:113926. doi: 10.1016/j.jviromet.2020.113926

17. Hirotsu Y, Omata M. Detection of R.1 lineage severe acute respiratory syndrome coronavirus 2 (SARS-CoV-2) with spike protein W152L/E484K/G769V mutations in Japan. PLOS Pathogens. 2021;17(6):e1009619. doi: 10.1371/journal.ppat.1009619.

18. Hirotsu Y, Omata M. Discovery of a SARS-CoV-2 variant from the P.1 lineage harboring K417T/E484K/N501Y mutations in Kofu, Japan. Journal of Infection. 2021;82(6):276–316. doi: https://doi.org/10.1016/j.jinf.2021.03.013.

19. Hirotsu Y, Omata M. SARS-CoV-2 B.1.1.7 lineage rapidly spreads and replaces R.1 lineage in Japan: Serial and stationary observation in a community. Infection, Genetics and Evolution. 2021;95:105088. doi: https://doi.org/10.1016/j.meegid.2021.105088.

20. Shepard SS, Meno S, Bahl J, Wilson MM, Barnes J, Neuhaus E. Viral deep sequencing needs an adaptive approach: IRMA, the iterative refinement meta-assembler. BMC Genomics. 2016;17:708. doi: 10.1186/s12864-016-3030-6. PubMed PMID: 27595578; PubMed Central PMCID: PMCPMC5011931.

21. Hadfield J, Megill C, Bell SM, Huddleston J, Potter B, Callender C, et al. Nextstrain: real-time tracking of pathogen evolution. Bioinformatics. 2018;34(23):4121–3. doi: 10.1093/bioinformatics/bty407. PubMed PMID: 29790939; PubMed Central PMCID: PMCPMC6247931.

22. Rambaut A, Holmes EC, O’Toole A, Hill V, McCrone JT, Ruis C, et al. A dynamic nomenclature proposal for SARS-CoV-2 lineages to assist genomic epidemiology. Nat Microbiol. 2020;5(11):1403–7. doi: 10.1038/s41564-020-0770-5. PubMed PMID: 32669681.

23. Shu Y, McCauley J. GISAID: Global initiative on sharing all influenza data - from vision to reality. Euro Surveill. 2017;22(13). doi: 10.2807/1560-7917.ES.2017.22.13.30494. PubMed PMID: 28382917; PubMed Central PMCID: PMCPMC5388101.

24. Qassim SH, Chemaitelly H, Ayoub HH, AlMukdad S, Tang P, Hasan MR, et al. Effects of BA.1/BA.2 subvariant, vaccination, and prior infection on infectiousness of SARS-CoV-2 Omicron infections. medRxiv. 2022:2022.03.02.22271771. doi: 10.1101/2022.03.02.22271771.

25. Chadeau-Hyam M, Tang D, Eales O, Bodinier B, Wang H, Jonnerby J, et al. Omicron BA.1/BA.2 variant transition in the Swedish population reveals higher viral quantity in BA.2 casesThe Omicron SARS-CoV-2 epidemic in England during February 2022. medRxiv. 2022:2022.03.10.22272177. doi: 10.1101/2022.03.10.22272177.

26. Kirsebom FCM, Andrews N, Stowe J, Toffa S, Sachdeva R, Gallagher E, et al. COVID-19 Vaccine Effectiveness against the Omicron BA.2 variant in England. medRxiv. 2022:2022.03.22.22272691. doi: 10.1101/2022.03.22.22272691.

27. Emma B. Hodcroft. CoVariants: SARS-CoV-2 Mutations and Variants of Interest. 2021.

28. Stegger M, Edslev SM, Sieber RN, Cäcilia Ingham A, Ng KL, Tang M-HE, et al. Occurrence and significance of Omicron BA.1 infection followed by BA.2 reinfection. medRxiv. 2022:2022.02.19.22271112. doi: 10.1101/2022.02.19.22271112.

29. Baggio S, L’Huillier AG, Yerly S, Bellon M, Wagner N, Rohr M, et al. Severe Acute Respiratory Syndrome Coronavirus 2 (SARS-CoV-2) Viral Load in the Upper Respiratory Tract of Children and Adults With Early Acute Coronavirus Disease 2019 (COVID-19). Clinical Infectious Diseases. 2020;73(1):148–50. doi: 10.1093/cid/ciaa1157.

30. Colson P, Fournier P-E, Delerce J, Million M, Bedotto M, Houhamdi L, et al. Culture and identification of a “Deltamicron” SARS-CoV-2 in a three cases cluster in southern France. medRxiv. 2022:2022.03.03.22271812. doi: 10.1101/2022.03.03.22271812.

31. Madera S, Crawford E, Langelier C, Tran NK, Thornborrow E, Miller S, et al. Nasopharyngeal SARS-CoV-2 viral loads in young children do not differ significantly from those in older children and adults. Scientific Reports. 2021;11(1):3044. doi: 10.1038/s41598-021-81934-w.

32. Heald-Sargent T, Muller WJ, Zheng X, Rippe J, Patel AB, Kociolek LK. Age-Related Differences in Nasopharyngeal Severe Acute Respiratory Syndrome Coronavirus 2 (SARS-CoV-2) Levels in Patients With Mild to Moderate Coronavirus Disease 2019 (COVID-19). JAMA Pediatrics. 2020;174(9):902–3. doi: 10.1001/jamapediatrics.2020.3651.

33. Ochoa V, Díaz FE, Ramirez E, Fentini MC, Carobene M, Geffner J, et al. Infants Younger Than 6 Months Infected With SARS-CoV-2 Show the Highest Respiratory Viral Loads. The Journal of Infectious Diseases. 2021;225(3):392–5. doi: 10.1093/infdis/jiab577.

34. Jones TC, Biele G, Mühlemann B, Veith T, Schneider J, Beheim-Schwarzbach J, et al. Estimating infectiousness throughout SARS-CoV-2 infection course. Science. 2021;373(6551):eabi5273. doi: doi:10.1126/science.abi5273.

35. Paul LA, Daneman N, Schwartz KL, Science M, Brown KA, Whelan M, et al. Association of Age and Pediatric Household Transmission of SARS-CoV-2 Infection. JAMA Pediatrics. 2021;175(11):1151–8. doi: 10.1001/jamapediatrics.2021.2770.

36. Jackson B, Boni MF, Bull MJ, Colleran A, Colquhoun RM, Darby AC, et al. Generation and transmission of interlineage recombinants in the SARS-CoV-2 pandemic. Cell. 2021;184(20):5179-88.e8. doi: https://doi.org/10.1016/j.cell.2021.08.014.

37. Bolze A, Basler T, White S, Rossi AD, Wyman D, Roychoudhury P, et al. Evidence for SARS-CoV-2 Delta and Omicron co-infections and recombination. medRxiv. 2022:2022.03.09.22272113. doi: 10.1101/2022.03.09.22272113.

38. UK Health Security Agency. SARS-CoV-2 variants of concern and variants under investigation in England, Technical briefing 39. 2022.

